# Research protocol for a multidimensional environmental and health impact study of petrochemical plant emissions in Calvert city, Kentucky

**DOI:** 10.64898/2026.07.07.26356427

**Authors:** Luz Huntington-Moskos, Matthew Cave, Lark Reynolds, Lauren B. Anderson, Bradley Housman, Mikus Abolins-Abols, Rhonda Fratzke, Rochelle H. Holm, Ted Smith

**Author notes:** Corresponding author: Rochelle H. Holm, Center for Healthy Air Water and Soil, Christina Lee Brown Envirome Institute, School of Medicine, University of Louisville, 302 E. Muhammad Ali Blvd., Louisville, KY 40202, United States.

## Abstract

While exposure to volatile organic compounds such as ethylene dichloride and vinyl chloride monomer is a well-established cause of liver disease, particularly hepatic hemangiosarcoma, characterizing real-world exposure profiles in communities surrounding industrial centers remains challenging. Calvert City, Kentucky (population ∼2,500), provides a unique setting characterized by both active industrial emissions and legacy sources of air toxics. To address these complexities, this method paper describes the framework for the Biomonitoring and Environmental Assessment for Community Outreach and Neighborhood Safety (BEACON) study. By utilizing a novel, multi-dimensional exposure assessment strategy, BEACON aims to characterize air toxic exposures and provide actionable data for community health and safety. For the BEACON study, we will leverage Kentucky Department of Air Quality measures of air toxics, analyze urine samples in a small cohort of community volunteers, analyze community urine via wastewater in an adjacent community, geocode citizen odor reporting, assess blood markers in wildlife, survey small and large animal veterinarians in the area for anomalies in morbidity and mortality, and work with the regional health system to enhance vigilance for health issues associated with toxicants present in the area. In addition, blood samples will be collected at three time points and biobanked for future analyses. Efforts will be made to link this study to additional large-scale long-term cohorts where possible. Throughout the project, community engagement will play a critical role by raising awareness, fostering collaboration, and ensuring that the voices of affected residents are heard.

## 1. INTRODUCTION

Exposure to ethylene dichloride (EDC) and vinyl chloride monomer (VCM) is associated with documented health risks, particularly hepatotoxicity.^1–3^ Individuals residing near a VCM/Polyvinyl Chloride (PVC) production plant have been reported to have higher urinary concentrations of thiodiglycolic acid (TdGA), a metabolite of VCM and EDC.^3^ Elevated TdGA levels have been associated with an increased risk of liver fibrosis. Vinyl chloride is known to undergo metabolic activation in the liver, leading to the formation of reactive intermediates that can induce DNA damage and promote carcinogenesis.^2,4^ Hepatic metabolism converts lipophilic substances into water-soluble metabolites for urinary excretion; the concentration of these analytes in urine and municipal wastewater provides a measurable index of population-wide chemical exposure.^5,6^ Hemangiosarcoma, an extremely rare and aggressive vascular tumor, has been strongly associated with exposure to vinyl chloride^7,8^ and related volatile organic compounds (VOC).^9^ The mechanism by which vinyl chloride contributes to hepatic hemangiosarcoma may involve the disruption of normal endothelial cell function and the promotion of angiogenesis, both of which are fundamental to the pathogenesis of vascular tumors.^10^ Furthermore, the latency period for the development of hepatic hemangiosarcoma can vary, further complicating the identification of additional cases. These findings underscore the potential impacts of EDC and VCM emissions from petrochemical industries on liver health among nearby populations.

Calvert City, Kentucky, is a small rural town (population 2,517) characterized by overlapping current and legacy sources of industrial contamination. Ongoing emissions from the Calvert City Industrial Complex, including VOCs (such as EDC), together with a history of industrial accidents, coexist with legacy contamination from former unlined waste ponds, industrial landfills, and hazardous-waste management facilities, some of which are listed on the U.S. Environmental Protection Agency (EPA) National Priority List (NPL) of Superfund sites. These sources have resulted in persistent impacts on air quality, soil, groundwater, and adjacent surface waters, with implications for chronic community exposure through multiple environmental pathways. Community members have long expressed concerns about repeated exposure to toxic chemicals, including Group 1 carcinogens EDC, vinyl chloride, and benzene.^11,12^ These chemicals emissions have been reported to have broad carcinogenic potential including an increased risk of breast cancer,^13^ as well as rare cancers such as angiosarcoma, with multiple cases documented in the area.^14^ The Kentucky Department of Air Quality (KDAQ) began collecting 24-hour ambient air concentrations of VOCs (EDC, vinyl chloride, 1,3-butadiene, acrylonitrile, and benzene) at three monitoring locations in 2020 as part of a special-purpose monitoring study conducted in cooperation with the U.S. EPA.^15^ The monitoring locations included a waste management facility (Calvert City Liquid Waste Disposal [LWD]; targeting maximum EDC concentration), Johnson-Riley Road (targeting maximum vinyl chloride concentration), and Calvert City Elementary School (targeting air quality in more heavily populated areas). In December 2024, a fire at a petrochemical plant in Calvert City resulted in the release of EDC.^11^ This incident followed a history of similar events originating from the same plant.^12,16^ In response to ongoing environmental and public health concerns, the community has sought data to inform public health decision-making and environmental remedial action. The occurrence of multiple hemangiosarcoma cases within a relatively short period,^14^ combined with a history of unpermitted emissions and environmental consent decrees, raises concerns regarding broader potential public health risks.

The Biomonitoring and Environmental Assessment for Community Outreach and Neighborhood Safety (BEACON) study will be conducted to address the need for an integrated, community-engaged approach to characterize environmental exposures and strengthen public health preparedness. BEACON is designed as a multidimensional environmental health assessment that combines regulatory air-monitoring data, human biomonitoring, wastewater-based exposure assessment, community odor reporting, veterinary and wildlife surveillance, and regional health-system engagement. Specifically, the study will leverage KDAQ air toxics measurements collected using canister-based sampling; assess urinary biomarkers of exposure in a cohort of community volunteers; evaluate pooled community-level urinary exposure markers through wastewater sampling in a nearby community; geocode and analyze citizen-reported odors; conduct wildlife blood sampling; survey small- and large-animal veterinarians regarding unusual patterns of morbidity or mortality; and collaborate with the regional health system to enhance vigilance for health outcomes plausibly associated with chemical air exposures. In addition, blood samples will be collected from participants at three time points and biobanked to support future analyses as new hypotheses, biomarkers, and analytic methods emerge. The overarching goal of BEACON is to assess potential environmental and health impacts associated with chemical emissions and industrial incidents in Calvert City while establishing a rapid-response framework that can be activated during future industry-related or disaster-related exposure events. By integrating environmental measurements, biological markers, community observations, animal health indicators, and clinical awareness, the BEACON study seeks to provide a more comprehensive understanding of exposure pathways and support timely, transparent, communication with residents, public health agencies, regulators, clinicians, and industry stakeholders.

**Figure 1.**
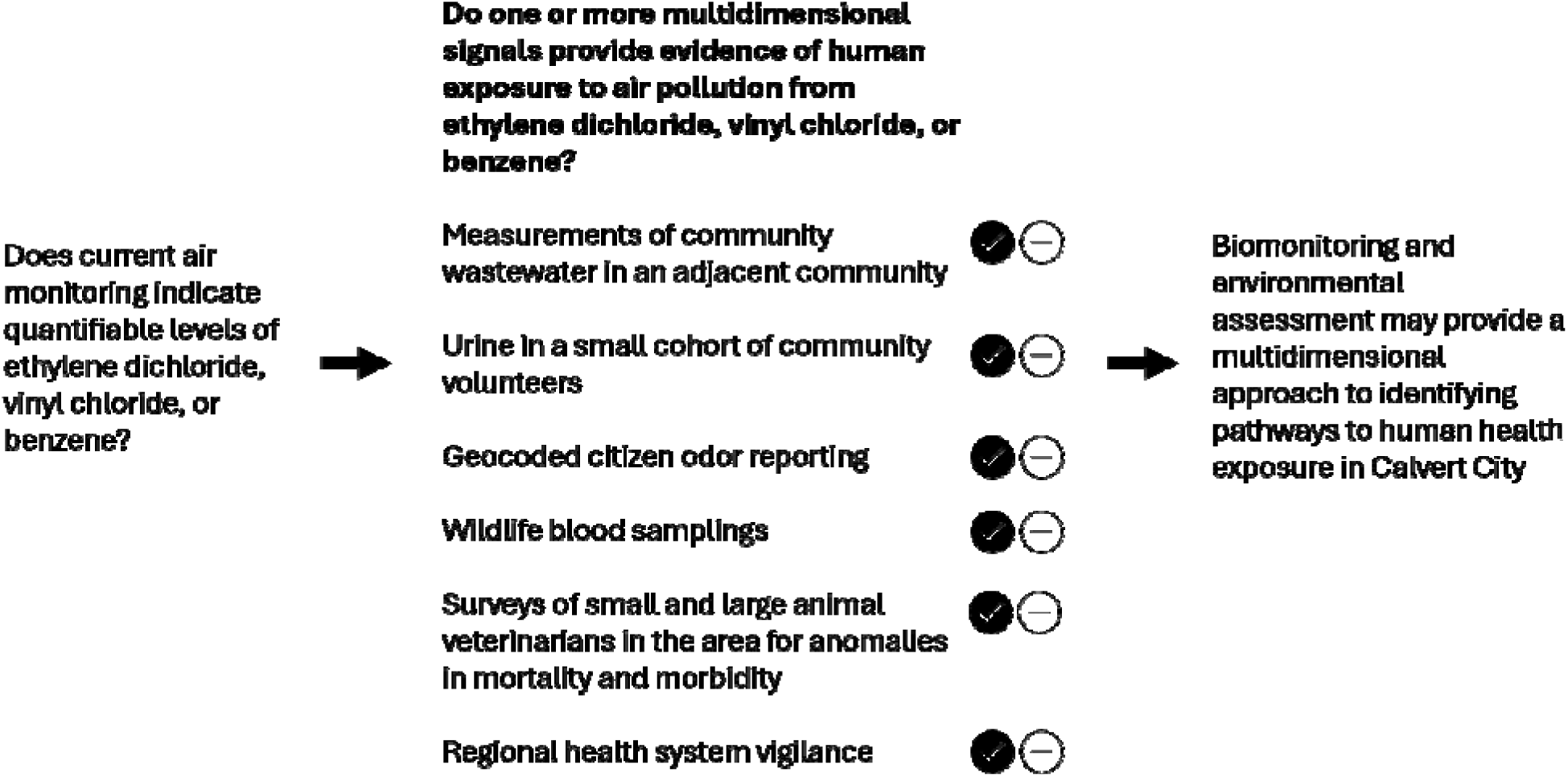
Conceptual model illustrating the convergent multidimensional measures incorporated into the Biomonitoring and Environmental Assessment for Community Outreach and Neighborhood Safety (BEACON) study to assess the environmental and health impacts of petrochemical plant emissions in Calvert City, Kentucky.

### 1.1 Research questions

The BEACON research protocol proposed in this study is designed to address the following research questions:

> Question 1: To what extent do current air-monitoring data demonstrate ongoing, quantifiable ambient exposure to ethylene dichloride, vinyl chloride, and benzene in the Calvert City study area, and how do concentrations vary across sampling locations and over time?

This research is intended to establish baseline ambient concentrations of hazardous air pollutants over a one-year monitoring period. Calvert City has experienced recurring accidents and VOC emissions, with ambient concentrations of some VOCs among the highest reported in the nation.^15^ Between October 2020 and December 2021, the KDAQ conducted canister air-capture monitoring in Calvert City at six-day intervals.^17^ The aim of that study was to estimate 24-h average concentrations of VOCs from passive canister samples to assess cancer risk. Concentrations were compared with national VOC data from the Air Quality System database. EDC and benzene exceeded their respective chronic screening levels at all locations, with Calvert City monitors recording the highest levels nationally.^15^ Vinyl chloride exceeded screening levels at three locations, while 1,3-butadiene exceeded at two. At the Johnson-Riley Road monitoring site, the cumulative cancer risk was estimated to be 1×10 , indicating a potential for 100 additional cancer cases per one million individuals exposed continuously over a 70-year lifetime. At the LWD monitoring site, the estimated cumulative cancer risk was 1×10 ³, corresponding to approximately 1,000 additional cancer cases per one million individuals exposed over the same period.^15^ The 2024 KDAQ air monitor data continues to show persistent elevated VOC concentrations in Calvert City (Table 1). Given these efforts, our study aims to contribute to the quantifiable evidence to improve the understanding of ongoing environmental health exposure risks within the community.

> Question 2: Do convergent exposure indicators provide complementary evidence of ongoing VOC exposure concerns in the Calvert City study area, and do these measures add interpretive value beyond ambient air-monitoring data alone? Convergent exposure indicators include urinary metabolites of ethylene dichloride, vinyl chloride, benzene, and related VOC air toxics in a cohort of community volunteers; wastewater-based measures of pooled community urinary metabolites in an adjacent community; geocoded citizen odor reports; wildlife blood sampling; veterinarian-reported anomalies in small- and large-animal morbidity or mortality; and regional health vigilance.

**Table 1:** Kentucky Division of Air Quality (KDAQ) 2024 daily mean ambient concentrations of 1,3-butadiene, acrylonitrile, benzene, ethylene dichloride, and vinyl chloride, at Calvert City, Kentucky.

| Parameter | Daily mean (Parts per billion Carbon) |  |  |
| --- | --- | --- | --- |
|  | Calvert City<br>Elementary | Johnson-Riley<br>Road | LWD |
| 1,3-Butadiene | 0.022 | 0.023 | 0.047 |
| Acrylonitrile | 0.005 | 0.000 | 0.002 |
| Benzene | 0.740 | 0.900 | 0.845 |
| Ethylene dichloride | 0.271 | 0.553 | 6.378 |
| Vinyl chloride | 0.069 | 1.072 | 1.012 |

Characterizing community health risk requires distinguishing among sources, exposure pathways, and exposures. In the Source–Pathway–Exposure framework, industrial emissions and legacy hazardous waste sites represent potential sources; ambient air represents a primary exposure pathway through which the community gets exposed to air pollutants; and biological or population-level measurements provide evidence that exposure has occurred. Air monitoring can therefore determine whether VOCs are present in the ambient environment and whether inhalation represents a plausible exposure pathway. However, air monitoring alone does not establish the extent to which contaminants are reaching the broader community. For this reason, BEACON integrates ambient air monitoring with convergent exposure measures, including urinary metabolites in community volunteers, wastewater-based measures of pooled urinary metabolites, wildlife blood sampling, geocoded odor reports, veterinary surveillance, and regional health vigilance. The degree to which these exposure indicators align spatially or temporally with air-monitoring results remains poorly characterized and is therefore a central focus of this study.

The December 2024 accident occurred at a petrochemical facility with a documented history of recurring fugitive emissions affecting the surrounding community.^11^ Members of the Calvert City community contacted the University of Louisville Center for Integrative Environmental Health Sciences Community Engagement Core (CIEHS) to request further investigation of the potential health risks associated with environmental exposures in their community. This community concern, combined with the identification of these rare cancer cases,^14^ underscores the need for immediate and comprehensive assessment of potential environmental exposures and their associated health risks in the surrounding community. The present study provides an opportunity to examine the additive value of a multi-dimensional approach to characterize exposure to air toxics by integrating environmental monitoring, biomonitoring, and complementary exposure indicators.

### 1.2 Research objectives

This study has the following seven objectives. Each objective provides contemporaneous measurements across multiple environmental, biological, and community-based factors.

Objective 1: Analyze KDAQ air monitoring data to assess and quantify levels of EDC, vinyl chloride, benzene, and other hazardous air pollutants over a one-year monitoring period.

Objective 2: Establish a baseline for wastewater surveillance within the wastewater catchment serving a community adjacent to Calvert City, Kentucky, by identifying metabolites indicative of pooled community exposures on a weekly basis over a one-year period.

Objective 3: Use a community-engaged approach to identify 25 community members for monthly urine sample collection to measure exposure biomarkers and assess liver function, quarterly symptom tracking, and collect and biobank blood samples.

Objective 4: Use the Smell MyCity app to identify geographic and temporal areas of concern over a one-year period.

Objective 5: Evaluate the utility of wild birds as sentinel species for VOC exposure by assessing bird health markers from Calvert City and control areas.

Objective 6: Survey regional veterinarians to assess reports of morbidity, mortality, and unusual clinical findings in small and large animals as potential early indicators of community exposure to VOCs.

Objective 7: Engage with the regional health system and healthcare providers, including oncologists, to enhance vigilance for health issues associated with exposures to petrochemical plant emissions.

## 2. MATERIALS AND METHODS

### 2.1 Overview of study design

The study is motivated by historical and recent industrial release events, the presence of air toxics of toxicological concern, and the need for a rapid-response framework that can be applied to future industrial or disaster-related exposure events. Rather than relying on a single environmental or biological measure, the study adopts an integrated Source–Pathway–Exposure framework to distinguish potential emission sources, environmental transport through ambient air, and measurable indicators of exposure in humans, wastewater, animals, and community-reported observations. The study will generate coordinated environmental, biomonitoring, and health-context data to improve interpretation of current exposure conditions and to support evidence-based public health decision-making. Specifically, these data will be used to assess whether measured ambient air toxic concentrations correspond with convergent exposure indicators, identify conditions under which exposure-reduction strategies may be warranted, and inform community, clinical, and policy responses to industrial emissions. If convergence is observed across these complementary data streams, the findings may also inform the development of more timely, scalable, and cost-effective approaches for exposure screening and public health response in other small rural communities facing similar environmental health concerns.

### 2.2 Study setting

Calvert City, Kentucky, in Marshall County (Figure 1), is a small industrial community along the Tennessee River where residential areas coexist with a large chemical manufacturing complex consisting of several chemical manufacturing industries. The area is characterized by both legacy contamination and ongoing industrial emissions. A major legacy source is the B.F. Goodrich Superfund Site,^18^ where remediation remains ongoing under U.S. EPA oversight. In addition, the area continues to be affected by active chemical operations, some of which have been subject to federal and state consent decrees related to environmental compliance. Calvert City is home to the largest U.S. industrial emitter of EDC, a key intermediate in vinyl chloride production, and is characterized by the potential for cumulative community exposure to multiple VOC air toxics, including EDC, vinyl chloride, benzene, and 1,3-butadiene, emitted from three chemical manufacturing facilities.

**Figure 1.**
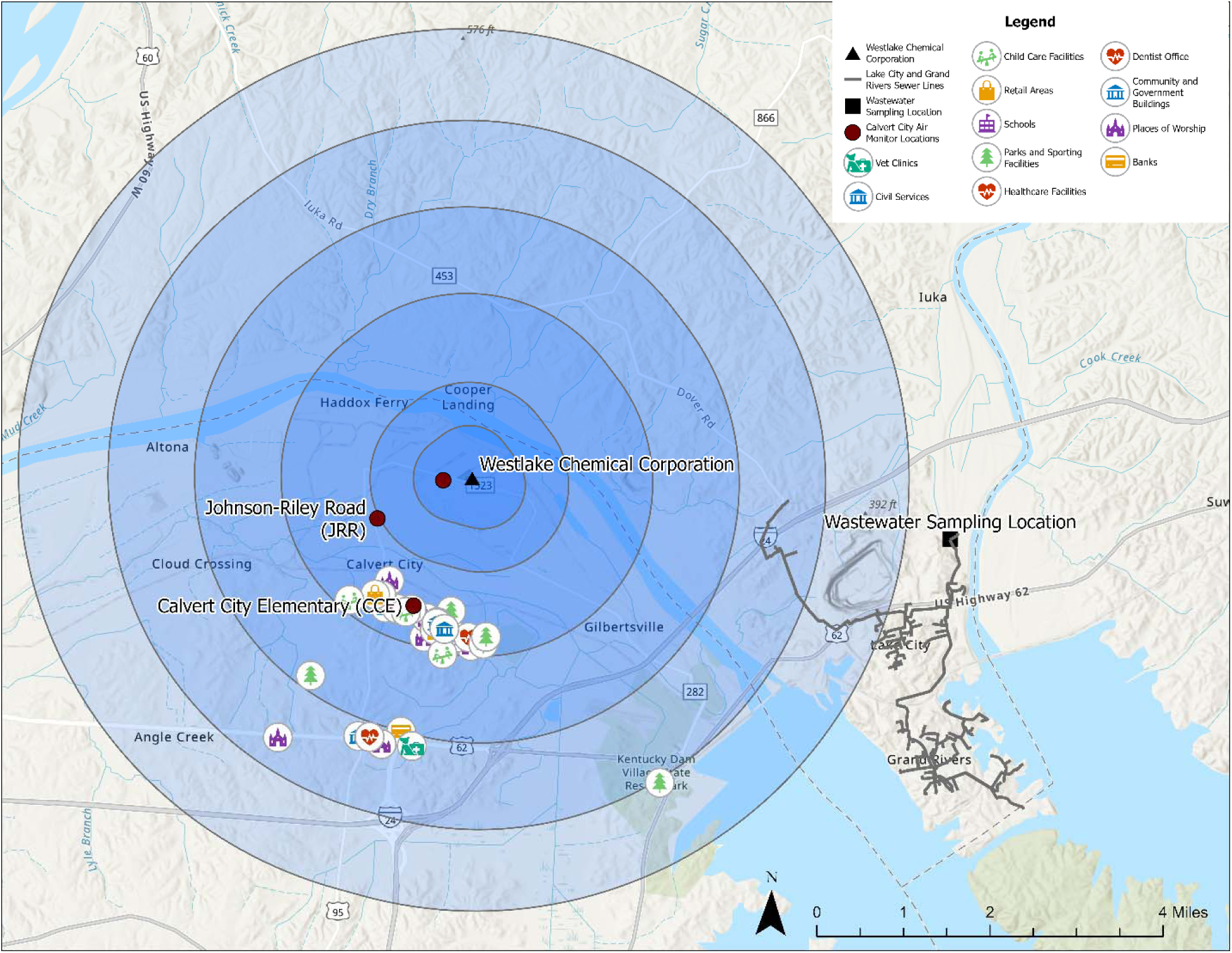
Study area in Calvert City, Kentucky. Locations of the air monitoring and wastewater sampling sites, as well as the community sites of interest, are indicated.

### 2.3 Eligibility and enrollment

Air chemistry is monitored on a 1-in-6-day sampling schedule by the KDAQ. Site selection, sampling design, and data collection are managed by the KDAQ for Calvert City.^17^

A medical monitoring program will recruit 25 adult, voluntary, community participants through snowball recruitment strategies. The study will 1) gather monthly urine samples over 12 months for investigating the metabolites of VOCs, 2) track symptoms of exposure through a quarterly symptom tracking survey, 3) collect blood samples at three time points, and 4) develop and provide an “rapid response” self-collection urine sample kit in case of exposure events. Early in Year 1, the research team conducted a community meeting to review study goals, share information on methods, and discuss scheduling for biosample collection to allow community driven inclusion in medical monitoring program eligibility and enrollment.

Given concerns about the potential occurrence of additional hemangiosarcoma cases, blood collection will build on previous metabolomic research conducted in Louisville among vinyl chloride–exposed polyvinyl chloride workers by extending this approach to evaluate candidate biomarkers and biological pathways implicated in the development of hepatic hemangiosarcoma within the wider community.^8,19^ These biomarkers will include bile acids, gamma-glutamyl amino acids, and fibrin degradation products.

The urine and wastewater samples are convergent, both investigating the metabolites of VOCs and using the same laboratory and analytical method. We established a wastewater monitoring program to demonstrate the feasibility of passive, population-based, assessment of exposure to air toxics by collecting sewage samples from a community 5.5 miles east of Calvert City for one year. The nearby location was selected because the project was unable to obtain cooperation from the municipal government where the industrial complex is located. Wastewater reflects a community chemical quantification of urinary metabolites.^6^ Enrollment is therefore anonymous. Weekly, a 125 mL aliquot of a 24-hour composite wastewater sample will be transferred into a polyethylene terephthalate bottle, stored on ice during sampling and transportation, and delivered to the University of Louisville for analysis.^20,21^ The samples will be stored at −80°C until analysis using previously described methods.^6^

In addition to the medical monitoring, the research team has collaborated with the Southern Environmental Health Study (SEHS) based at Vanderbilt University. This SEHS cohort study, established in 2022, will enroll 35,000 study participants in southern states from Delaware to Texas and including Kentucky. The goal of SEHS is to understand the relationship between environmental exposures and cancer outcomes in the Southern United States. At the 3- and 6-month biosample collection visits, the research team will offer BEACON study participants the opportunity to connect with SEHS and provide them with a silicone wristband. When worn by an individual, the silicone wristband serves as a measurement tool that passively absorbs chemical exposures. The silicone wristband will be worn for seven days and nights continuously. After wearing the silicone bands for seven days and nights, the participant will complete a 10-item paper survey regarding their experience wearing the wristband and then, place and seal the wristband in the provided silver bag. Subsequently, the sealed silver bag containing the wristband and the completed paper questionnaire will be placed in the prepaid envelope provided and mailed to the SEHS research team at Vanderbilt. The wristbands will be analyzed by the SEHS research team.

The Smell MyCity app^22^ is a community-based reporting tool that enables residents to document odor events in real time, including information on smell type, intensity, location, and associated symptoms. Many VOCs and other air toxics have recognizable odor signatures, and odor events may occur intermittently, during short-duration releases, or under meteorological conditions that are not fully captured by fixed-site or time-limited air-monitoring networks. Although odor reports cannot identify or quantify specific contaminants and should not be interpreted as direct chemical concentrations, they provide a novel, community-sensed complement to conventional air monitoring by capturing the timing, location, perceived character, and associated symptoms of odor episodes as experienced by residents. These crowd-sourced data will be aggregated and analyzed to identify odor hotspots, temporal patterns, potential overlap with air-monitoring results, industrial activity, meteorological conditions, and community health concerns. As part of the study, the Smell MyCity app will be promoted within the study area to encourage participants and other community members to document odors encountered in their daily environments. Because the app is publicly available and can be used anywhere in the United States, this approach provides an opportunity to capture broader community observations that may help contextualize local patterns of perceived air quality and support the project’s efforts to better understand the exposure pathways. Smell MyCity data are anonymous, and no personal identifiers are collected. Fact sheets will be developed to describe the functionality of the Smell MyCity app and provide step-by-step instructions for downloading and using the app. These materials will be distributed during community meetings and through electronic folders to increase accessibility. A short, publicly available instructional video will also be developed to facilitate app download and use.

The wildlife health component of BEACON is informed by recent work from Abolins-Abols and colleagues^23^ demonstrating that urban birds can provide spatially resolved biological signals of environmental conditions. In Louisville, Kentucky, Abolins-Abols et al.^23^ integrated high-resolution estimates of nitrogen dioxide, ultrafine particles, and vegetation with intensive sampling of American robins and found that higher nitrogen dioxide exposure within 500 m of capture locations was associated with elevated baseline corticosterone, a glucocorticoid marker of physiological stress.^23^ Related recent work from the Abolins-Abols group has extended this sentinel-species approach by evaluating erythrocyte indices as potential markers of vascular stress in adult American robins^24^ and micronuclei and other nuclear abnormalities as markers of genotoxic damage in robin blood cells.^25^ Together, these studies support the rationale for using common songbirds as organism-level indicators that may complement environmental monitoring and help characterize local biological responses to air pollution. In Calvert City, the wildlife health study will evaluate whether wild birds show biological evidence consistent with environmental exposure, physiological stress, or cellular injury. The team will collaborate with local landowners and state agencies to access private and state property in and around the study area. Using approved wildlife research protocols, permitted mist-net capture methods, and blood sampling procedures conducted under all applicable state and federal wildlife research licenses, the research team will capture representative songbird species present in the study area. Blood-based measurements will be used to assess candidate markers of oxidative stress, hematologic or cellular injury, genotoxic damage, and potential toxicant exposure. Wildlife data will be interpreted in conjunction with air-monitoring results and other BEACON exposure indicators and will not be used as standalone evidence of emission source attribution, disease causation, or human health risk.

The veterinary surveillance component of BEACON is grounded in a One Health and animal-sentinel framework, which recognizes that human, animal, and environmental health are interconnected and that companion animals, livestock, and wildlife may provide complementary information about shared environmental conditions.^26–29^ Animals can serve as valuable sentinel species when they occupy the same geographic area as human populations, share relevant environmental media, have behaviors that increase their potential for contaminant exposure, or exhibit measurable clinical, pathologic, or biomarker responses that warrant further investigation. This approach is particularly relevant in settings with complex, intermittent, or spatially heterogeneous exposures, where fixed environmental measurements may not fully reflect biologically meaningful contact with contaminants over time. In the BEACON study, the research team will engage regional veterinarians serving the Calvert City area, including practitioners caring for companion animals, horses, and livestock, to identify patterns of animal illness, unusual diagnoses, and premature mortality that may warrant further investigation. The study will include outreach to veterinarians and structured interviews to identify conditions of concern, including unexplained premature mortality, hepatobiliary disease, hematologic or vascular abnormalities, and histologically documented vascular sarcoma–type cancers, including hepatic angiosarcoma or hemangiosarcoma when documented in veterinary records. Veterinary observations will be evaluated as exploratory, hypothesis-generating sentinel information and interpreted in conjunction with ambient air monitoring, human biomonitoring, wastewater-based measurements, odor reports, and wildlife biomarker data. These data will not be used as standalone evidence of source attribution, disease causation, or direct human health risk. Rather, they will provide an additional community-level perspective for assessing whether animal health observations converge spatially or temporally with other BEACON exposure indicators.

Baptist Health Paducah will serve as the regional clinical and health-system partner for BEACON study. As a major regional medical and referral center, Baptist Health Paducah serves approximately 200,000 patients annually across a four-state area through its 373-bed hospital and a network of more than 45 care locations.^30^ Its service region includes communities in western Kentucky, southern Illinois, and surrounding areas, including Marshall County and Calvert City. Baptist Health Paducah also operates the Ray & Kay Eckstein Regional Cancer Care Center, which is identified in the hospital’s Community Health Needs Assessment as the region’s only cancer center, with medical oncology, radiation oncology, chemotherapy and infusion, diagnostic and screening, and surgical services. This role is especially relevant because Baptist Health Paducah serves a region with documented environmental health concerns related to both chemical manufacturing and legacy radiological and chemical contamination. This includes reported VOC air-toxic concerns in Calvert City, as well as contamination at the Paducah Gaseous Diffusion Plant Superfund site involving technetium-99, VOCs, PCBs, and other hazardous contaminants.^15,31^ Through BEACON, Baptist Health Paducah will be provided with environmental, biomonitoring, and population-level findings to support provider education, enhance awareness among oncology and primary care, inform community education, and, where supported by the findings, guide consideration of appropriate screening or clinical monitoring strategies for exposure-associated conditions, including hemangiosarcoma and related vascular sarcoma–type cancers.

### 2.4 Outcome measures: Environmental and clinical

Air chemistry data will be obtained from the KDAQ ambient air monitoring program through publicly available U.S. EPA data. Air chemistry is monitored by the KDAQ on a 1-in-6-day sampling schedule using a Xonteck Model 911a sampler with 6-Liter stainless steel canister. Twenty-four-hour integrated ambient air samples are collected to measure hazardous air pollutant concentrations and establish community air toxics levels. These monitoring data are submitted to U.S. EPA, where they are made publicly available for analysis. KDAQ conducts canister-based metered air monitoring through a capture system coupled with gas chromatography-mass spectrometry (GC-MS) analysis^32^. It employs EPA Method TO-15A, the standardized method for measuring volatile organic compounds in ambient air using 24-hour composite capture. The monitoring locations of greatest relevance are Johnson-Riley Road and the LWD superfund site (Figure 2), which have previously indicated elevated risks for cancer. These data will be used alongside community wastewater data, recruited participant urinary metabolites, animal data, and healthcare reports to examine contemporaneous patterns across exposure media.

Wastewater and recruited participant urinary metabolites measurements will be based on a unified liquid chromatography-mass spectrometric approach combining targeted and untargeted analyses for community exposure.^6^ The targeted assay will quantify 29 established VOC biomarkers. The untargeted assay will measure 818 Phase II metabolites (239 mercapturic acids, 431 glucuronides, and 148 sulfates) from 145 parent xenobiotics, enabling detection of both known and putative biotransformation products. These data will be compared with other source findings to provide a more comprehensive assessment of environmental exposures and associated health risks.

Our research team will recruit participants, gather clinical data, and provide phlebotomy expertise to support this work. A baseline survey will be completed prior to biological sample collection and will include standardized demographic questions including sex, home address, work address, occupational history, etc. Urine samples will be analyzed by the University of Louisville. Blood samples will be biobanked at the CIEHS Translational Research Support Core. An open-access, electronic folder will be made available to enrolled participants containing slides from previous community meetings, an overview of the BEACON study, information on the CIEHS, and a factsheet on Smell MyCityapp and its use. When used by community members, the Smell MyCity app will collect time-stamped and location-based odor reports submitted by residents when they experience unusual or unpleasant odors through a smartphone or web interface. The qualitative data collected through the Smell MyCity app will be categorized based on odor detail (Table 2). By combining community observations with environmental and health data, Smell MyCity will help document residents’ experiences of odors when research team members are not present and provide complementary data to support public health analyses and inform policy decisions.

**Table 2.** Code analysis for reported odors from the Smell MyCity app.

| **ID** | **Resident-reported odor detail** |
| --- | --- |
| 1 | Biological Mixture (fecal, rancid, decay/death) |
| 2 | Chemical Mixture (plastic, glue, rubber, sweet, polish remover, burnt/burning, garlic, cat pee/ammonia/urine, acid, industrial, chemical, alcohol) |
| 3 | Natural Gas/Fuel (gas, oil, fuel, cabbage, rotten eggs, sulfur) |
| 4 | Sewage |
| 5 | Misc. (garbage, trash, open burns, drugs, etc.) |

For the wildlife health pilot, blood sample measurements will include assessing the potential formation of micronuclei, quantifying relative telomere length, counting young versus old red blood cells, measuring DNA damage, and assessing oxidative stress markers. Upon capture, a small blood sample (200 µL, not exceeding 1 % of bird’s body weight) will be collected from the brachial vein using a 26 G needle in heparinized microcapillary tubes. A fraction of the blood sample will be immediately frozen on dry ice for DNA damage assays. Another fraction will be immediately centrifuged to separate plasma and erythrocytes and then frozen on dry ice for downstream analysis of blood redox state. Lastly, we will make blood smears for downstream cell counts and nuclear abnormality tests. Following blood collection, we will collect standard morphometric measurements from each bird and release them at the capture location. Blood collection and body mass measurement protocols will follow the methods described by Abolins-Abols et al.^23^

The survey of regional veterinarians will be primarily qualitative and will include case counts by animal type and reported symptoms that may be associated with air pollution exposure.

Baptist Health plans to use the data gathered to evaluate a geographic pattern of patients with conditions of interest and to identify areas that may warrant further investigation. These findings will inform retrospective reviews of clinical records to assess whether the occurrence of selected conditions differ across geographic areas with varying potential for environmental exposure. In collaboration with the oncology department, the findings will help guide our efforts in community and provider education, as well as clinical awareness.

### 2.5 Sample size

The sampling approach varies with each component of this multidimensional project. Air sampling will be conducted at two locations on a 7-day interval for the first month, followed by a monthly sampling over a one-year period. Wastewater sampling will be conducted at a single location on a weekly basis over a one-year period. A feasibility pilot involving 25 community members will be conducted to establish the biomonitoring component of the study and to provide preliminary measurements of exposure biomarkers and liver function. Smell MyCity data collection will be performed for one year, collecting as many reports as possible. Wildlife health pilot measurements and the survey of regional veterinarians will be conducted over a 1-year period. Healthcare system objectives will be implemented over a 1-year period; the sample size will be determined by community participation and provider engagement, with recruitment conducted to the maximum extent possible.

### 2.6 Analysis

The wastewater analysis results will be compared with ambient air monitoring data for concurrence and trends over time, using the concentrations of each VOC measured through air monitoring at each corresponding time point. The levels will be summarized using the mean and standard deviation and compared with the corresponding cancer unit risk estimates.

Descriptive statistics will be used to summarize baseline survey findings. For monthly urine biomarkers, mixed linear models will be applied to examine the association between each biomarker and those measured through air monitoring and wastewater surveillance. Generalized linear mixed-effects models will be employed to assess whether urine biomarkers are associated with liver function, using data from the 25 community members. The biobanked blood specimens will be analyzed at a later time.

Smell MyCity reports will be analyzed to identify areas where industrial and chemical odor reports cluster. ArcGIS Pro (Redlands, CA) Kernel Density tool will be used to identify areas with high density of noxious odor reports across the study area.

For the wildlife study, generalized linear mixed-effects models will be employed to assess the relationship between bird health metrics, distance from Calvert City Industrial Complex, and KDAQ air quality data. We will also verify if these relationships differ across bird species.

Veterinary chart review data will be analyzed using descriptive epidemiologic methods to characterize patterns in animal morbidity, mortality, age at death, and diagnoses of conditions of concern, including liver angiosarcoma-type cancers, across the study region. Analyses may include summary statistics, stratification by species, animal type (e.g., pets, livestock, and wildlife), clinic service area, and time period; and, where sample size permits, exploratory comparisons to identify unusual frequencies, temporal clustering, or geographic patterns that may warrant additional environmental health investigation.

Regional health system will engage with various care providers, including oncologists, to enhance vigilance for health issues associated with these exposures and to support appropriate clinical evaluation and analysis.

As data related to air toxics, wastewater surveillance, biospecimens and animals become available, researchers will meet with community members to assist in contextualizing the data presented in aggregate. Once the data are discussed, the research team will work in partnership with community members to decide next steps, which may include educational campaigns, public meetings, and outreach programs to support data sharing (community-level report-back^33^) and support informed decision-making. During the initial year of community discussions and air monitoring, information will be shared regarding the University of Louisville’s long history with vinyl chloride research,^19^ wastewater-based epidemiology,^20,21,34^ and the ways in which these research efforts have helped empower community leaders to improve public health. The discussion and co-designing of protocols will involve a review of standardized and relevant resources from the PhenX Toolkit^35^, including standardized items related to sex, gender, race/ethnicity, occupational history, and residence, as well as standardized protocols for blood draws, blood pressure measurements, and urine collection.

Throughout the study, community engagement will play a critical role, with efforts focused on raising awareness, fostering collaboration, and ensuring that the voices of affected residents are heard. As data are collected and analyzed, the research team will provide community members with both individual-level and community-level report-back of the research findings. With individual-level report-back, the research team will offer supportive consultation to assist community members in interpreting data for which no established clinical guidelines exist. The community-level report-back will provide community members with the opportunity to share context for the research findings and to discuss potential action steps and future research priorities. Depending on the community priorities, the next steps may include educational campaigns, public meetings, and outreach programs. This two-year project will begin with the establishment of environmental monitoring systems and initial clinical recruitment and data collection. The second year will focus on data analyses to ascertain strategies that are most likely to reduce cancer and cardiometabolic risk in the community, while further validating the wastewater surveillance and subclinical metabolomics associated with exposure. By combining robust scientific analysis with community-engaged action, this project aims to improve understanding of environmental health challenges faced by fenceline communities and lay the groundwork for policies that reduce health risks.

## 3. LIMITATIONS

This multidimensional approach offers a promising framework for evaluating environmental exposures and their potential health impacts; however, several limitations should be considered. The reduced air sampling frequency after the first month may overlook short-term pollution spikes. Additionally, with only two air study monitoring locations, the study may not fully capture the variability in community exposure. Meteorological factors such as wind patterns may further complicate data interpretation, potentially leading to misrepresentation of pollutant levels at points in time. Although limited KDAQ air study monitoring locations exist, the use of the Smell MyCity app may provide additional data points to capture potential changes in air quality.

Wastewater surveillance, while valuable for population-level exposure assessment, also introduces challenges, including public concerns over privacy. Additionally, wastewater measurements reflect not only community exposure but may also capture occupational exposure as workers return home to the adjoining community. Associations between air and wastewater pollutant levels may be complex due to differences in the timing of ambient air measurements and wastewater measurements. The results from the urine samples and biobanked blood samples are not diagnostic; as a result, they cannot be used to diagnose any disease or condition. Further, there are no established guidelines for safe levels of these chemicals in the body. The small sample size limits the generalizability of the findings but will provide valuable information on the feasibility and scalability of this approach.

Also, community members may be lost-to-follow up after the initial community meeting due to busy schedules or outside commitments. Therefore, we will survey participant regarding the days and times that are most convenient for group meetings and will provide a virtual meeting option for those who are unable to leave their home on a scheduled meeting date. For community members who do not have access to smartphone technology or WIFI, the research team will provide access to the technology, including a mobile hotspot for internet access. We will keep community members informed on research timelines to support a timely report-back (both at the individual and community level) and support continued engagement with the project. As such, we will discuss aspects of report-back at community meetings using aggregated data to ensure 1) participant privacy and 2) that the report-back format meets community needs and addresses community concerns.

Ongoing community engagement is essential to build trust, encourage bidirectional communication, and facilitate the implementation of methods to be used in this multidimensional project. If modifications to the study are required, established community engagement will allow the BEACON research team to respond effectively to the community needs and emerging exposure events.

## Notes

The authors declare no competing financial interest.

## Data Availability

There is no data produced in the present study.

## References

(1) Cave, M.; Falkner, K. C.; Ray, M.; Joshi-Barve, S.; Brock, G.; Khan, R.; Bon Homme, M.; McClain, C. J. Toxicant-associated steatohepatitis in vinyl chloride workers. Hepatology 2010, 51 (2), 474–481. DOI: 10.1002/hep.23321

(2) U.S. Environmental Protection Agency. IRIS Toxicological Review of Vinyl Chloride (Final Report). 2020. https://iris.epa.gov/Document/&deid=232144

(3) Yuan, T. H.; Chen, J. L.; Shie, R. H.; Yeh, Y. P.; Chen, Y. H.; Chan, C. C. Liver fibrosis associated with potential vinyl chloride and ethylene dichloride exposure from the petrochemical industry. Science of The Total Environment 2020, 739, 139920. DOI: 10.1016/j.scitotenv.2020.139920

(4) Wang, W.; Qiu, Y.; Jiao, J.; Liu, J.; Ji, F.; Miao, W.; Zhu, Y.; Xia, Z. Genotoxicity in vinyl chloride-exposed workers and its implication for occupational exposure limit. . American Journal of Industrial Medicine 2011, 54 (10), 800–810. DOI: 10.1002/ajim.20990.

(5) Kumar, R.; Adhikari, S.; Driver, E. M.; Smith, T.; Bhatnagar, A.; Lorkiewicz, P. K.; Xie, Z.; Hoetker, J. D.; Halden, R. U. Towards a novel application of wastewater-based epidemiology in population-wide assessment of exposure to volatile organic compounds. Science of The Total Environment 2022, 845, 157008. DOI: 10.1016/j.scitotenv.2022.157008

(6) Lorkiewicz, P.; Chen, J. Y.; Hoetker, D.; Talley, D.; Anderson, L. B.; Dusterhoff, A.; Holm, R. H.; Smith, T.; Srivastava, S. A unified liquid chromatography-mass spectrometric approach combining targeted and untargeted analyses for community exposure profiling in wastewater. Environment International 2026, 212, 110293. DOI: 10.1016/j.envint.2026.110293.

(7) Makk, L.; Creech, J. L.; Whelan, J. G.; Johnson, M. N. Liver damage and angiosarcoma in vinyl chloride workers: A systematic detection program. Jama 1974, 7;230(1), 64–68.

(8) Guardiola, J. J.; Hardesty, J. E.; Beier, J. I.; Prough, R. A.; McClain, C. J.; Cave, M. C. Plasma metabolomics analysis of polyvinyl chloride workers identifies altered processes and candidate biomarkers for hepatic hemangiosarcoma and its development. Int J Mol Sci 2021, 22 (10). DOI: 10.3390/ijms22105093

(9) Carreon, T.; Hein, M. J.; Hanley, K. W.; Viet, S. M.; Ruder, A. M. Coronary artery disease and cancer mortality in a cohort of workers exposed to vinyl chloride, carbon disulfide, rotating shift work, and o-toluidine at a chemical manufacturing plant. Am J Ind Med 2014, 57 (4), 398–411. DOI: 10.1002/ajim.22299.

(10) Fujiwara, R. Exposure to sub parts per million levels of vinyl chloride can increase the risk of developing liver injury. Hepatology Communications 2018, 2 (3), 227–229. DOI: 10.1002/hep4.1169.

(11) Youngblood, J.; Watkins, M. Burn at Westlake Vinyls Plant sparks fears of toxic chemicalrelease. 2024. https://www.wpsdlocal6.com/news/burn-at-westlake-vinyls-plant-sparks-fears-of-toxic-chemicalrelease/article_066bcd68-c7c2-11ef-9ff0-7b072f9dc0c6.html.

(12) Youngblood, J.; Patterson, E. Lawsuit over chemical emissions from Westlake Vinyls couldbe dismissed. 2024. https://www.wpsdlocal6.com/news/lawsuit-over-chemical-emissions-from-westlake-vinyls-could-bedismissed/article_87937a2e-8a75-11ef-b1e9-7baf41c470d4.html.

(13) Heck, J.; He, D.; Wing, S.; Ritz, B.; Carey, C.; Yang, J.; Stram, D.; Le Marchand, L.; Park, S.; Cheng, I.;, et al. Exposure to outdoor ambient air toxics and risk of breast cancer: The multiethnic cohort. Int J Hyg Environ Health 2024, 259, 114362. DOI: 10.1016/j.ijheh.2024.114362.

(14) Song, L. The polluter just got a million-dollar fine. that won’t cure this woman’s rare cancer. 2022. https://www.propublica.org/article/pollution-kentucky-westlake-chemical-cancer

(15) U.S. Environmental Protection Agency. Calvert City, Kentucky Volatile Organic Compound (VOC) Air Quality Risk Assessment. 2024.

(16) Operle, D. Calvert City families file suit against Westlake Vinyls for gross negligence after EPA report. WKMS, 2024. https://www.wkms.org/environment/2024-02-07/calvert-city-families-file-suit-against-westlake-vinyls-for-gross-negligence-after-epa-report.

(17) U.S. Environmental Protection Agency. Calvert City, Kentucky Air Monitoring. 2024. https://www.epa.gov/ky/calvert-city-kentucky-air-monitoring (accessed 2025 January 5).

(18) U.S. Environmental Protection Agency. *B.F. Goodrich*. 2026. https://www.epa.gov/superfund-redevelopment/superfund-sites-reuse-kentucky

(19) Falk, H.; Creech, J. L.; Heath, C. W.; Johnson, M. N.; Key, M. M. Hepatic Disease Among Workers at a Vinyl Chloride Polymerization Plant. JAMA 1974, 230, 59–63.

(20) Yeager, R.; Holm, R. H.; Saurabh, K.; Fuqua, J. L.; Talley, D.; Bhatnagar, A.; Smith, T. Wastewater sample site selection to estimate geographically resolved community prevalence of COVID-19: A sampling protocol perspective. Geohealth 2021, 5 (7), e2021GH000420. DOI: 10.1029/2021GH000420.

(21) Holm, R. H.; Mukherjee, A.; Rai, J. P.; Yeager, R. A.; Talley, D.; Rai, S. N.; Bhatnagar, A.; Smith, T. SARS-CoV-2 RNA abundance in wastewater as a function of distinct urban sewershed size. Environmental Science: Water Research & Technology 2022, 8 (4), 807–819. DOI: 10.1039/d1ew00672j.

(22) Rangel, A.; Anderson, L.; Holm, R. H.; Smith, T. Resident odor reports and differing health outcomes in areas of high industrial emission odor, Louisville, Kentucky. *Progress in Community Health Partnerships: Research*, Education, and Action 2026, 20 (2), 271–276.

(23) Abolins-Abols, M.; Yeager, R.; Turner, J.; Smith, T.; Bhatnagar, A. Greenness and pollution exposure predict corticosterone concentration in an urban songbird. Frontiers in Physiology 2025, 16, 1603811. DOI: 10.3389/fphys.2025.1603811.

(24) Stine, L. A.; Abolins-Abols, M. The impact of air pollution on the cardiovascular system in American Robins: NO2 levels’ and ultrafine particulate levels’ correlation to the ratio of immature to mature erythrocytes. The Cardinal Edge 2024, 2(2).

(25) Mittlestat, E. Micronuclei as a marker of genotoxicity in an urban songbird [Senior honors thesis, University of Louisville]. ThinkIR: The University of Louisville’s Institutional Repository 2025.

(26) National Research Council. Animals as sentinels of environmental health hazards; National Academies Press, 1991. DOI: 10.17226/1351.

(27) Rabinowitz, P. M.; Gordon, Z.; Holmes, R.; Taylor, B.; Wilcox, M.; Chudnov, D.; Nadkarni, P.; Dein, F. J. Animals as sentinels of human environmental health hazards: an evidence-based analysis. EcoHealth 2005, 2 (1), 26–37. DOI: 10.1007/s10393-004-0151-1.

(28) Reif, J. S. Animal sentinels for environmental and public health. Public Health Reports 2011, 126. DOI: 10.1177/00333549111260S108.

(29) van der Schalie, W. H., Gardner, H. S., Jr, Bantle, J. A., De Rosa, C. T., Finch, R. A., Reif, J. S., Reuter, R. H., Backer, L. C., Burger, J., Folmar, L. C., & Stokes, W. S. Animals as sentinels of human health hazards of environmental chemicals. Environmental Health Perspectives 107(*4*), 309–315.

(30) Baptist Health Paducah. 2026. https://www.baptisthealth.com/locations/baptist-health-paducah

(31) U.S. Environmental Protection Agency. Paducah gaseous diffusion plant (USDOE) Kevil, KY. 2026.

(32) McGlenny, W. A.; Pleil, J. D.; Evans, G. F.; Oliver, K. D.; Holdren, M. W.; Winberry, W. T. Canister-based method for monitoring toxic VOCs in ambient air. Journal of the Air & Waste Management Association 1991, 41 (10), 1308–1318.

(33) Korfmacher, K. S.; Brody, J. G. Moving forward with reporting back individual environmental health research results. Environmental Health Perspectives 2023, 131 (12), 125002. DOI: 10.1289/EHP12463

(34) Holm, R. H.; Osborne Jelks, N.; Schneider, R.; Smith, T. Beyond COVID-19: Designing Inclusive Public Health Surveillance by Including Wastewater Monitoring. Health Equity 2023, 7 (1), 377–379. DOI: 10.1089/heq.2022.0055.

(35) Hamilton, C. M.; Strader, L. C.; Pratt, J. G.; Maiese, D.; Hendershot, T.; Kwok, R. K.; Hammond, J. A.; Huggins, W.; Jackman, D.; Pan, H.;, et al. The PhenX Toolkit: get the most from your measures. American Journal of Epidemiology 2011, 174 (3), 253–260. DOI: 10.1093/aje/kwr193.

